# Dynamics of mask use as a prevention strategy against SARS-Cov-2 in Panama

**DOI:** 10.1101/2021.09.15.21263479

**Authors:** Hermógenes Fernández-Marín, Gaspar Bruner-Montero, Ana Portugal-Loayza, Virginia Miranda, Alcibíades Villareal, Eduardo Ortega, Virginia Núñez-Samudio, Iván Landires, Luis C. Mejia, Sandra López-Vergès, William T. Wcislo, K.S. Jagannatha Rao

## Abstract

**Background:** Early in the SARS-CoV-2 pandemic, many national public health authorities implemented non-pharmaceutical interventions to mitigate disease outbreaks. Panamá established mandatory mask use two months after its first documented case. Initial compliance was high, but diverse masks were used in public areas. We studied behavioral dynamics of mask use through the first two COVID waves in Panama, to improve implementation of effective, low-cost public health containment measures, when populations are exposed to novel air-borne pathogens.

**Methods:** Mask use behavior was recorded from pedestrians in four Panamanian populations (August to December 2020). We recorded facial coverings; and if used, the type of mask, and gender and estimated age of the wearer.

**Findings:** People were highly compliant (> 95%) with mask mandates, and demonstrated important population-level behaviours: 1) decreasing use of cloth masks over time, and increasing use of surgical masks; 2) mask use was 3-fold lower in sub-urban neighborhoods than other public areas; and 3) young people were least likely to wear masks.

**Interpretation:** Results help focus highly-effective, low-cost, public health interventions for managing and controlling a pandemic. Considerations of behavioural preferences for different masks, relative to pricing and availability, are essential for optimizing public health policies. Policies to increase availability of effective masks, and behavioral nudges to increase acceptance, and to facilitate mask usage, during the on-going SARS-CoV-2 pandemic, and for future pandemics of respiratory pathogens, are key tools, especially for nations lagging in access to expensive vaccines and pharmacological approaches.

**Funding:** 11-2020 SNI Grant, SENACYT.

## Introduction

During December 2019, an acute respiratory disease known as Coronavirus disease (COVID-19) was detected in the Chinese city of Wuhan, and the causative agent of the outbreak was named severe acute respiratory syndrome coronavirus 2 (SARS-CoV-2). SARS-CoV-2 has affected the public health systems of every country in the world (1), infecting more than 200 million people and causing over 4.3 million deaths since reported at December 2019 through August 2021. The COVID-19 pandemic has forced world-leading health organizations to provide recommendations on how public health systems should operate generally across the globe (2–3). A recommendation to minimize the transmission of the virus, for which the main transmission mechanism is via aerosol droplets (4–5), was the use of personal protective equipment (PPE) to protect healthcare workers and infected people. Not surprisingly, many governments adopted these recommendations, and further included mask usage for the general population with the integration of other non- pharmacological interventions to prevent the rapid transmission of the virus in the population.

Throughout the first year of the pandemic, the global public health system lacked the pharmacological tools for the prophylactic and therapeutic management of COVID-19. However, the implementation of non-pharmacological public health measures has allowed the containment, management, and control of the disease to varying degrees. Non- pharmacological measures, such as social distancing, self-isolation (including quarantine), changes in hygienic behavior (e.g., increased frequency of hand washing), and face-covering in public areas were preventive strategies suggested by international health agencies. Perhaps the use of face masks has been the most controversial strategy of prevention. There are many types of masks, and where, when, and how to use them can be complicated to evaluate, and for some individuals, mask use is perceived to impair individual freedom.

Face-covering of the general population was quickly adopted and recommended as one of the most important elements for preventing the spread of COVID-19 (6–8). The use of a facial mask was recommended because the social distance among persons (more than 1.5 meters) was difficult to maintain in indoor settings and crowded areas, and particularly given the high prevalence of presymptomatic and non-symptomatic cases. More importantly, the use of a facial mask may reduce the aerosol transmission of the virus from infected and non- symptomatic patients to the healthy population (8). Furthermore, high public compliance with mask usage is an effective and low-cost collective action that reduces viral transmission at the public and community level (9, 10). However the prevalence of different types of masks on the population and their different levels of protection can affect the impact of the transmission (8).

There is limited information about the prevalence of different types of masks at the population level (8–11). No study has reported how the usage of different types of masks has changed throughout the COVID-19 pandemic, or previous pandemics (10–11). Understanding behavioural responses to the received information about mask efficacy as well as to the supply and demand of masks during the pandemic can improve the efficiency of the system, facilitating transport logistics, mitigating the volatility of wholesale prices, and reducing the shortage of supplies between the general population and healthcare workers (12–15). Here we describe the behaviour of mask wearing during the end of the first wave and the beginning of the second wave of SARS-CoV2 infections in Panama.

## Materials and Methods

### Data collection

Observations of the type of mask used were recorded in urban areas of four provinces of the Republic of Panama, including the Provinces of Panama (Amelia Denis de Icaza in San Miguelito’s districts, and Felipillo (Pacora) at Panama’s districts), Cocle (city of Penonome), Veraguas (city of Santiago), and Chiriqui (city of David), between August 28th and December 12th, 2020. In each locality, data were recorded 4 to 6 days per month. Each day the observations were conducted in each one of the following places from August to December 2020: i) from 7:00 am to 8:00 am in bus and train terminals, ii) from 8:30 to 9:30 along public walkways of main avenues, iii) and from 10:00 to 11:00 am in supermarkets. From November to December 2020, additional data were collected in neighborhood areas from 4:00 pm to 5:00 pm. The data were recorded as i) biological gender (man or woman), ii) use of mask (correct or incorrect), iii) type of mask wearing (cloth, surgical, KN95, valve, or other types of masks, which included all type of masks that were not aforementioned), iv) the estimated range of the age (early adulthood < 19, middle adulthood 20 to < 40, late-middle adulthood 40 to < 60, and late adulthood > 60 (16–17). Masks were classified as follows: cloth masks are any fabric masks with cotton or synthetic cloth; surgical masks are dispensable colored non-medical masks of 3 layers; KN95 masks are non-medical masks of four or five layers curved design with contour adapted to the face; valved masks are considered any type of masks that used an exhalation valve; and the “others” category included any other facial covering, including scarves and kerchiefs. Initial observations in August and September indicated that some people wore masks incorrectly, which was tabulated as missing data. In October, November, and December, we scored mask use as “correct” or not, with the former defined as the mask covered the nose and mouth of the observed person; otherwise, it was tabulated as incorrect use. The price of masks in the market was quoted in Panama Compra (https://www.panamacompra.gob.pa) and local commercial shops, such as hardware stores, warehouses, pharmacies, supermarkets, and shopping malls.

### Data processing and analyses

To evaluate face mask usage (correct or incorrect) in public areas, we used a Generalized Linear Mixed Model (GLMM) with a binomial distribution and logic link function. After removing the missing data from 64 650 observations, the GLMM model was performed with 36 441 observations. The model included the predictor variables gender (male and female), place (main streets, market, neighborhood, and terminal station), and age (early, middle, late-middle, and late adulthood), and the interactions between gender × place, and gender × age. Region was included as a random effect to account for differences among populations, with the predictor variable place nested within region. A total of 64 650 observations were documented during the study.

We conducted all statistical analyses in R (www.r-project.org) (18). The GLMMs were generated using the ‘glmmTMB()’ function in the ‘glmmTMB’ package (19). The model selection was based on the Akaike Information Criteria comparing all models via the ‘AICtab()’ function in the ‘bbmle’ package (20). To validate our model, a residual diagnostic analysis was performed by simulating 1 000 times the model’s residuals using the ‘simulateResiduals()’ function in the DHARMa package (21). Overdispersion was tested with the ‘overdisp_fun()’ function and multicollinearity with the ‘check_collinearity()’ function in the ‘performance’ package. We used the Wald χ^2^ tests with a type III sum of squares to estimate the significant effects of our model using the ‘Anova()’ function in the ‘car’ package (22), and odds ratios (OR) were subtracted by exponentiating the coefficients of the model.

To determine whether the frequency of use of different types of masks changed over time, we used a multinomial logistic regression model (MLRM), which can help to characterize observations when the response variable has multiple categories. Mask type (cloth, KN95, surgical, valve, and others) was evaluated as a function of gender, place, and time period (continuous predictor variable) in the MLRM using the ‘multinom’ function in the *nnet* package (23). The MLRM was performed with 64 006 observations after removing missing data and variable “other masks”. Model selection, multicollinearity, and ORs were estimated as mentioned above. All graphs were generated with the function ‘ggplot()’ in the ‘ggplot2’ package (24), and final editions were performed in the program Inkscape (www.inkscape.org).

## Results

To evaluate face mask usage in the population, we analyzed 36 266 observations in different public areas. Among these observations, 34 266 (94%) people were observed wearing a mask, and 2 175 (6%) people were not wearing a mask. Men (93%, n = 20 122) and women (95%, n = 16 319) did not differ in the use of mask (GLMM, χ^2^ = 1·04, p=0·306). In contrast, face mask use differed at different places (GLMM, χ^2^ = 29·19, p < 0·0001): mask use was more prevalent among people at transport terminals (94 %, n = 15 810), main streets (95·1%, n = 13 894), and supermarkets (98·3%, n = 4 763), decreasing to 75% (n = 1 974) in neighborhood areas. Also, mask use differed with age (GLMM, χ^2^ = 18·94, p = 0·0002), used more frequently by middle (93·5%, n = 19 442), late-middle (95%, n = 11 673), and elderly ages (95.6%, n = 3 134), relative to young adults (90%, n = 2 192).

The odds ratios of people wearing masks between supermarkets, transport terminals station, and main streets did not differ (p > 0·05), but people in sub-urban neighborhood areas were 20 times (OR 0·05 [95% CI 0·01–0·16], p < 0·0001, figure 1) less likely to wear a mask. While the odd ratios of middle, late-middle, and late adulthood groups did not differ in face mask-wearing (p > 0·05), people in the early adulthood group are 1.49 times (OR 0·67 [95% CI 0·54–0·84], p < 0·0001, figure 1) less likely to wear a mask. For the interactions, there were no differences in odds between women and men at supermarkets and main streets areas (p > 0·05). In contrast, women at neighborhood areas (OR 1·78 [95% CI 1·34–2·38], p < 0·0001, figure 2A) and transport terminals (OR 1·46 [95% CI 1·18–1·81], p = 0·001, figure 2A), respectively, had a 78% and 56% increase in the odds of wearing a mask compared to men (figure 2B). While women in early, late-middle, and late adulthood groups were more likely to wear a mask than men (figure 2B).

**Figure 1.**
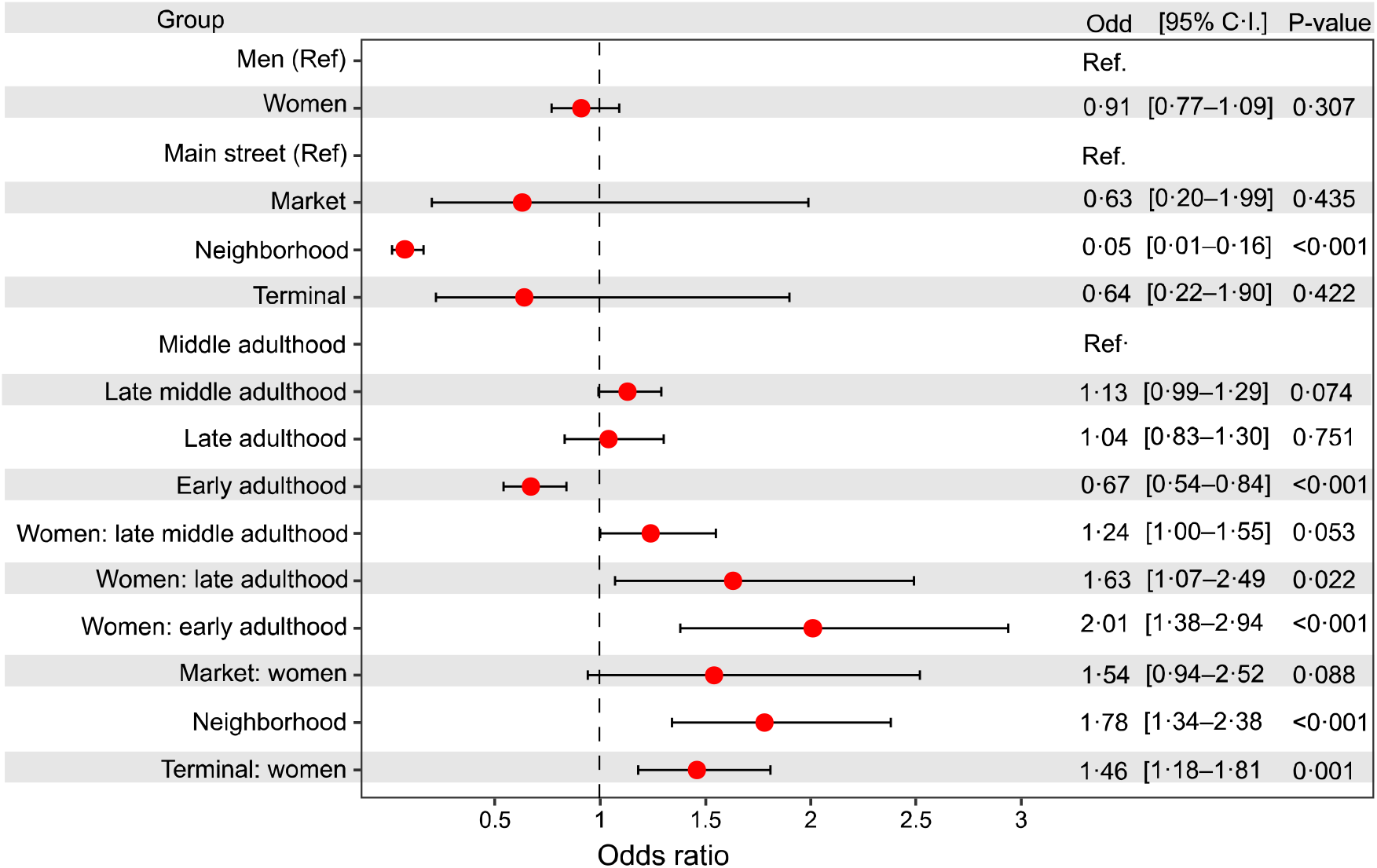
Odds ratios for people wearing a mask during the COVID-19 pandemic. Odds ratios and 95% confidence intervals for the group terms gender (men and women), place (mean street, market, neighborhood, and terminal), and age (early, middle, late-middle, and late adulthood). We included the interactions gender × place, and gender × age using a GLMM with binomial distribution, accounting for the random effect of region, and place nested within region. There were 36 441 observations included in the model. P-values denote the statistical significance of the model. Men, main streets, and middle adulthood were used as reference term within their groups denoted as Ref. The vertical dashed line represents the null value (odds ratio=1·0).

**Figure 2.**
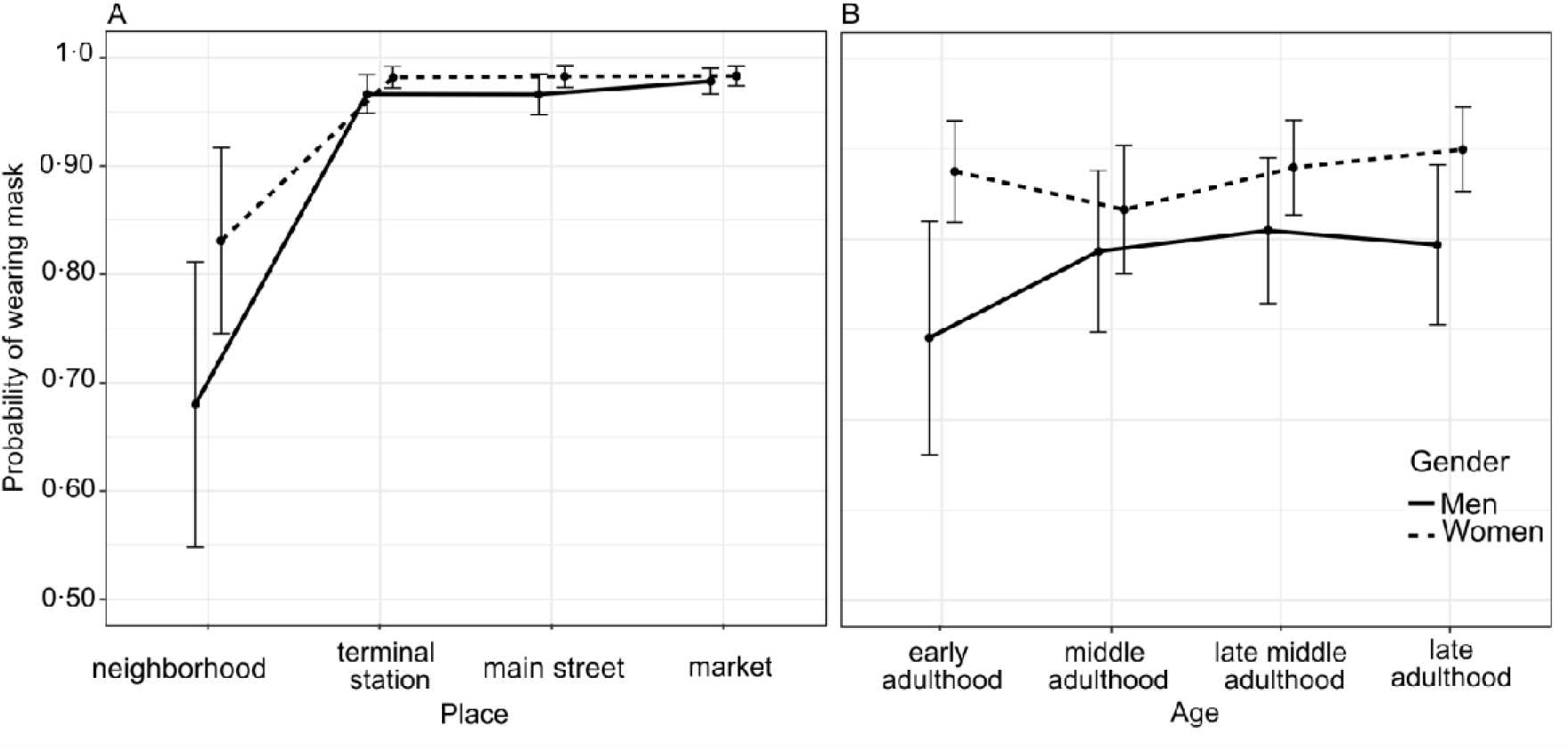
Wearing mask probability during the COVID-19 pandemic. (A) Interaction plot showing the probability of face mask-wearing between genders at different public areas, and (B) the probability of wearing masks between gender at different ages. The probabilities were estimated from a binomial GLMM across observations (n = 36 441). Error bars represent the standard error of the mean.

Different types of face mask protection were recorded (n = 64 650) from four areas of Panama (figure 3) during a rapid rise period of the COVID-19 pandemic (figure 4A). We found substantial differences in the use of difference types of masks in the population, with most people wearing surgical (68%, n = 44 184) and cloth (27.2%, n = 17 627) masks, and fewer people wearing KN95 (2.36%, n = 1527), valve masks (1.16%, n = 735), and other masks (0.89%, n = 577). Women were 3.5 times more likely to wear cloth (OR 3·54 [95% CI 2·96–4·25], p < 0·0001), KN95 (OR 3·61 [95% CI 2·94–4·44], p < 0·001), and surgical (OR 3·71 [95% CI 3·10–4·44], p < 0·001) masks than men, relative to people wearing valve masks (table 1). There was a significant decrease in people wearing cloth masks over time (OR 0·98 [95% CI 0·96–0·99], p = 0·001), figure 4B). Conversely, the odds of people wearing surgical masks over time increased 2% compared to people wearing valve masks (OR 1·02 [95% CI 1·01–1·04], p = 0·002), figure 4B). In neighborhood areas, people are 1·98 and 1·83 times more likely to wear cloth (OR 1·98 [95% CI 1·10–3·58], p = 0·023, table 1) and surgical (OR 1·83 [95% CI 1·02–3·29], p = 0·043, table 1) masks than valve masks, respectively. The late adulthood group is 1·53 and 2·17 times more likely of wearing cloth (OR 1·53 [95% CI 1·02– 3·29], p = 0·043) and KN95 (OR 2·17 [95% CI 1·60–2·94], p < 0·001) masks than valve masks (table 1).

**Table 1.** Odds ratio based on the multinomial logistic regression model.

| Response | Predictors | Odds ratio | 95% C.I. | Wald-test | P-value |
| --- | --- | --- | --- | --- | --- |
| Cloth vs valve |  |  |  |  |  |
|  | Women | 3.54 | 2.96 – 4.25 | 13.69 | **<0.001** |
|  | Time period | 0.98 | 0.96 – 0.99 | -3.39 | **0.001** |
|  | Market | 1.04 | 0.84 – 1.28 | 0.35 | 0.723 |
|  | neighborhood | 1.98 | 1.10 – 3.58 | 2.27 | **0.023** |
|  | Terminal | 1.1 | 0.93 – 1.31 | 1.14 | 0.253 |
|  | Early adulthood | 1.24 | 0.85 – 1.82 | 1.12 | 0.263 |
|  | Late middle adulthood | 1.01 | 0.86 – 1.18 | 0.09 | 0.931 |
|  | Late adulthood | 1.53 | 1.17 – 2.01 | 3.06 | **0.002** |
| KN95 vs valve |  |  |  |  |  |
|  | Women | 3.61 | 2.94 – 4.44 | 12.25 | **<0.001** |
|  | Time period | 1.02 | 1.00 – 1.03 | 1.9 | 0.057 |
|  | Market | 1.07 | 0.84 – 1.37 | 0.56 | 0.578 |
|  | neighborhood | 1.17 | 0.57 – 2.37 | 0.42 | 0.672 |
|  | Terminal | 0.9 | 0.74 – 1.10 | -1.01 | 0.312 |
|  | Early adulthood | 0.46 | 0.27 – 0.79 | -2.82 | **0.005** |
|  | Late middle adulthood | 1.12 | 0.93 – 1.36 | 1.17 | 0.242 |
|  | Late adulthood | 2.17 | 1.60 – 2.94 | 4.96 | **<0.001** |
| Surgical vs valve |  |  |  |  |  |
|  | Women | 3.71 | 3.10 – 4.44 | 14.3 | **<0.001** |
|  | Time period | 1.02 | 1.01 – 1.04 | 3.15 | **0.002** |
|  | Market | 0.96 | 0.78 – 1.18 | -0.4 | 0.69 |
|  | neighborhood | 1.83 | 1.02 – 3.29 | 2.02 | 0.043 |
|  | Terminal | 1.02 | 0.86 – 1.21 | 0.24 | 0.813 |
|  | Early adulthood | 0.95 | 0.65 – 1.38 | -0.29 | 0.773 |
|  | Late middle adulthood | 0.8 | 0.69 – 0.94 | -2.71 | **0.007** |
|  | Late adulthood | 1.19 | 0.91 – 1.56 | 1.27 | 0.204 |

Analysis was performed using a multinomial logistic regression model. The model included the response variable mask type (cloth, KN95, surgical, and valve) as a function of gender, place, and time period (continuous predictor variable).

**Figure 3.**
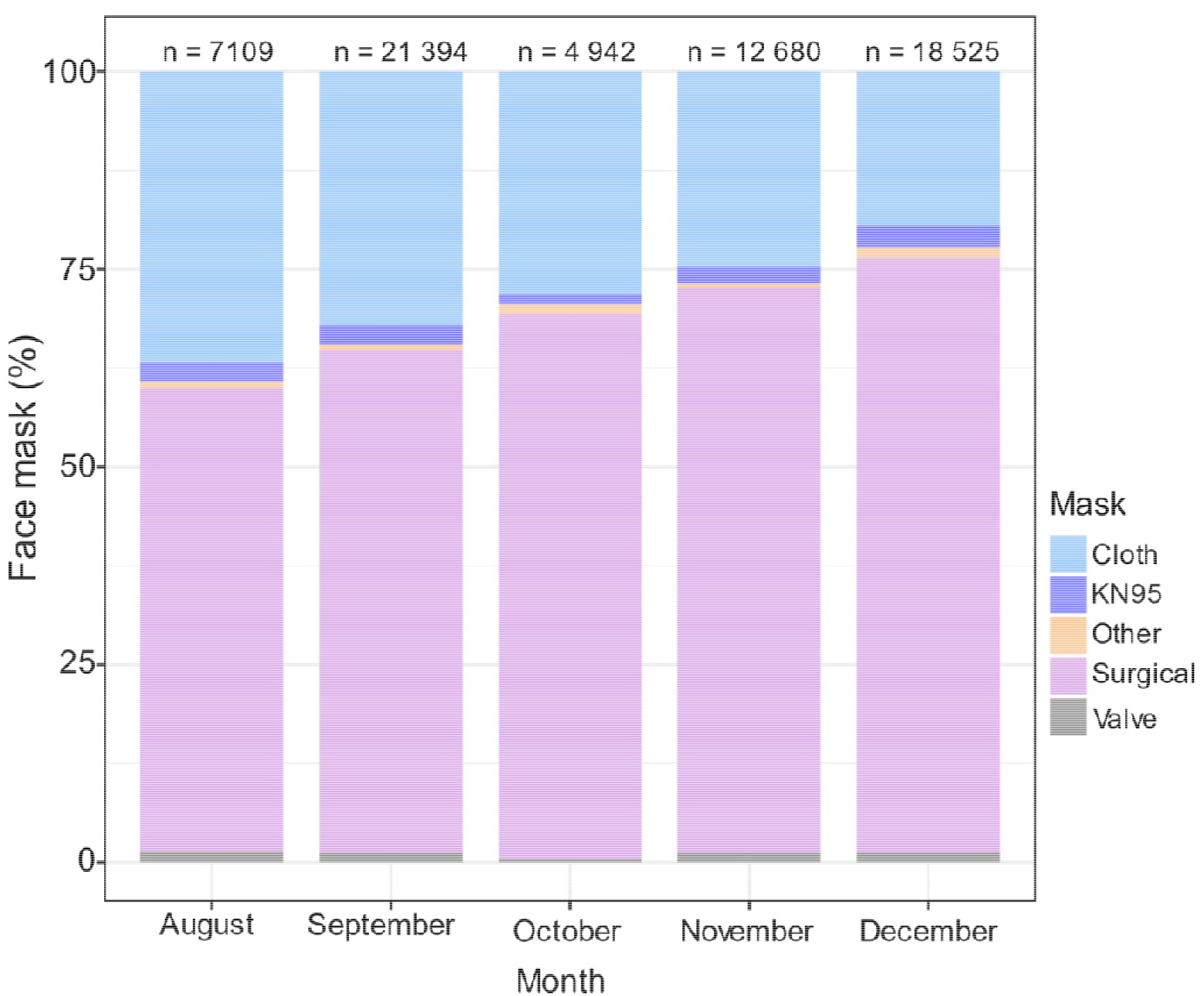
Usages of different types of face mask protection during the COVID-19 pandemic. Percentage of people wearing different types of masks in public areas between August and December 2020. A total of 64 650 observations were documented.

**Figure 4.**
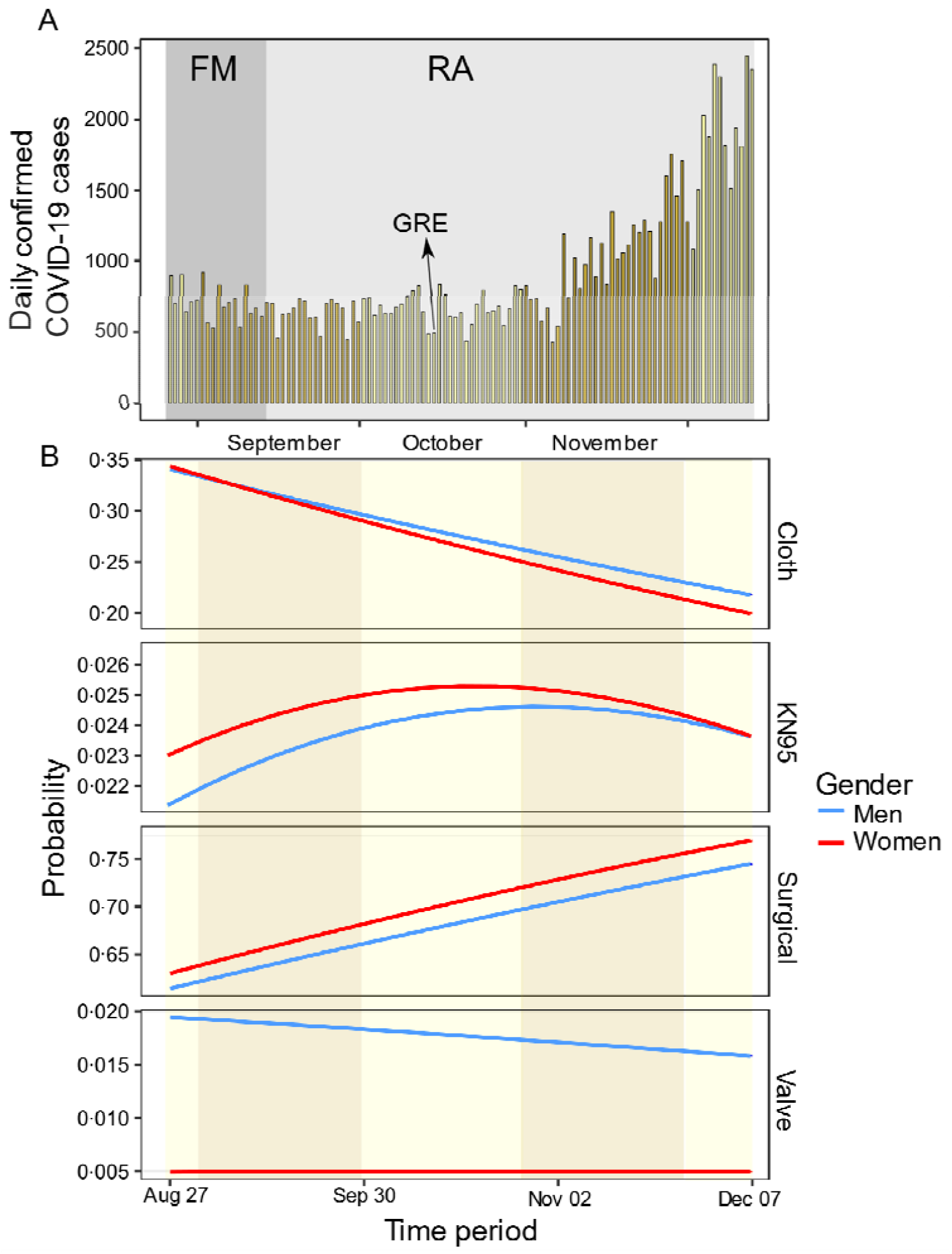
Different types of face mask protection over time during the COVID-19 pandemic. (A) Daily confirmed cases in Panama as a function of time across the observational study. (B) Multinomial logistic regression showing the predicted probability associated with the frequency of use for different types of face masks (cloth, KN95, surgical, and valve) in public areas over time. The model was performed with 64 006 observations, the categorical variable “other” was omitted from the model. A strict gender-based quarantine was mandated by the Panamanian government from April 1^st^, 2020 to February 2021, was lifted at different moments depending of the province and Rt. A relaxation of this policy with flexibility of movement (FM) in leaving homes is denoted by dark grey, and the reopening of economic activities is in light grey (RA); the arrow indicates the end of gender-based restrictions (GRE).

## Discussion

Panama is a developing country with a high income *per capita*, but with strong socioeconomic inequality such that a high percentage of the population live below the poverty line (25). The nation is a world-wide logistical and regional transportation hub, where more than 18 million people transit annually (26). Socioeconomic characteristics are not fundamentally different from many countries of the Latin American region. As pharmacological approaches are lagging in the region, due to global inequities in vaccine availability (10–11), encouraging hygienic prophylactic behaviour, mask usage and social distancing are key strategies to manage the COVID-19 pandemic. Panama reported its first positive cases of COVID-19 in early March 2020, and rapid community transmission was documented with a great diversity of strains (27). Severe containment and mobility-restriction measures were quickly implemented by the national government (27), reducing labor activity over 70%, with mandatory use of masks, enforced with fines. The prevalence and dynamics of mask use in Panama observed between August and December 2020, after the first and during the second epidemiological waves of SARS-CoV-2 positive cases, provide insights into the challenges faced and should facilitate preparedness for future pandemics.

The rapid spread of the COVID-19 pathogen created a huge demand world-wide for a limited supply of personal protective equipment (both for the general public and health care systems) and hospital equipment (respirators). The use of face masks during the first and second epidemiological SARS-CoV-2 waves in Panama showed a change in the behavior of the population in the use of types of masks. From August to December 2020, the use of surgical masks increased from 58% to 79%, while the use of cloth masks was reduced from 37% to 19%, while no changes were observed to the use of KN95 and valved masks in this period, both types of masks used by less than 3% of the people analyzed in this study. During the same period of time, a continuous decrease of the prices of non-medical surgical and KN95 masks was observed, whereas the price of cloth masks decreased only in December 2020 (table 2). Two non-competitive hypotheses can explain this change in strategy at the population level. The first hypothesis is the change in the purchasing power of people. Our data were taken in bus and subway terminals, and in main avenues in the different cities, which suggests that people with low and medium purchasing power were observed, so that work suspension could influence mask use at the population level. While a gradual economic recovery could change people’s purchasing power, enabling them to acquire surgical masks. Additionally, at the beginning of the pandemic cloth masks became part of a fashion culture, and producing them became a means of financial support for family businesses, but later information in the media and from public health authorities about the relative inefficiency of cloth masks, may have induced a decrease of their use as soon as people have learned that cloth masks did not offer them the best protection to prevent the spread of SARS-CoV-2 relative to surgical masks (10, 11). Surgical and KN95 particulate masks protect better from transmission by aerosols and large particles than fabric masks, but cloth masks protect better than no masks or improper use of them (11). However, it is possible that the knowledge about the protection efficacy conferred by the type of masks used was not reflected in an increased proportion of use of KN95 masks in the population, possibly because even with a decrease of its price, it remained 15 times more expensive than surgical masks. Subjectively, people report impressions that it was more difficult to breathe when using KN95 or N95 masks than surgical masks in the hot and humid weather in the tropics (28).

**Table 2.** Information about prices per unit of cloth, surgical and KN95 non-medical face masks selling in Panama from August to December 2020.

| Month (2020) | Prices per unit of surgical masks mean ± SD | Prices range per unit of cloth masks | Prices per unit of KN 95 masks mean ± SD |
| --- | --- | --- | --- |
| August | 0.19 ± 0.08 | 1.50-3.50 | 1.86 ± 0.95 |
| September | 0.08 ± 0.04 | 1.50-3.50 | 1.65 ± 0.97 |
| October | 0.09 ± 0.04 | 1.50-3.50 | 1.82 ± 0.91 |
| November | 0.07 ± 0.05 | 1.50-3.50 | 1.12 ± 0.28 |
| December | 0.07 ± 0.03 | 1.00-3.50 | 1.05 ± 0.88 |

Face mask prices in US\$ per unit (mean ± SD) or range. Prices were obtained from Panama Compra website (https://www.panamacompra.gob.pa/Inicio/#!/), and information from Instagram accounts of established commercial outlets.

Of relevance for public health strategies in reducing contagion rate is to identify elements in the behavior of the population that may shape dynamics of disease transmission, and then create policies around them to achieve public health objectives. Our study identified that people under the estimated age of 20, and at the neighborhood level, used masks less frequently. Patterns of mask use were similar among different sites in Panama, with lower compliance among the youth, which will be the last age group to be vaccinated, and should be targets of outreach and education, as they can be spreaders during the epidemic, especially with the origin and spread of new variants of SARS-CoV-2 like the Delta variant, with a higher risk to develop severe symptoms than at the beginning of the epidemic with the wild type virus. Scientific communication is crucial to keeping the population up-dated about the efficiency of the different strategies of prevention and the importance of protecting themselves, but also the role they could play in viral transmission and thus, the responsibility each person has in protecting others. Public health programs to make masks widely available, and mandate their use, represent highly effective, cost-efficient, solutions to decrease the spread of air-borne infectious diseases.

## Data Availability

Data sharing statements
Fernandez-Marin, Hermogenes (2021), Dynamics of mask use as a prevention strategy sgainst SARS-CoV-2 in Panama, Mendeley Data, V1, doi: 10.17632/92wp53s8sn.1

## Acknowledgments

We are grateful to Anna Melao for helping to collect some information on prices of masks. The Sistema Nacional de Investigación of the Republic of Panama granted funds to conduct the study. The study has the approval of the Ministry of Health and Bioethics permit from Comité Nacional de Bioética de la Investigación, Panamá.

## Author Contributions

Conceptualization: HFM, APL, VM; Study design and Methodology: HFM, APL; Literature search: HFM, JRKS, SLV, WTW; Data collection: HFM; Data analyses: GBM, HFM; Data interpretation: HFM, APL, VM, GBM, AV, LCM, SLV, WTW; Writing (first draft): HFM, GBM, APL, VM, AV, EO, VNS, IL, LCM, SLV, WTW, JRKS; Writing (Review and Editing): SLV, WTW; Funding acquisition: HFM. All authors have read and agreed to the published version of the manuscript.

## Data sharing statements

Fernandez-Marin, Hermogenes (2021), “Dynamics of mask use as a prevention strategy sgainst SARS-CoV-2 in Panama”, Mendeley Data, V1, doi: 10.17632/92wp53s8sn.1

## Compliance with Ethical Standards

An exemption review of our study was approved by Comité Nacional de Bioética de la Investigación de Panamá. All observations performed in this study not involving human participants, and was in accordance with the ethical standards, and the 1964 Helsinki declaration and its later amendments.

